# Nowcasting outbreak case and death counts from open-source reporting A validation on the 2026 Ebola (Bundibugyo) outbreak in the Democratic Republic of the Congo

**DOI:** 10.64898/2026.09.05.26362336

**Authors:** Rickard Nyman

## Abstract

Official outbreak counts are authoritative but published with delay. Between situation reports, a response team either works from a number that is several days old or extrapolates the official trend forward. We assess whether open-source reporting can reduce this gap. On the 2026 DRC Ebola outbreak we reconstruct the cumulative case and death curves from open sources, anchor them to the latest official figure, and project to the present. In a real-time (vintage) backtest over roughly ten weeks, using only reports available at each point, the open-source nowcast is comparable to simple extrapolation when the official figure is fresh, but once the official figure is stale by four to five days for cases, and about six for deaths, it is materially more accurate, cutting the median case-count error by about two thirds (12.6% to 3.9%) at a one-week lag. The advantage is concentrated at the phase transitions, where the official trend turns and any extrapolation of it is furthest from the truth: there the open-source nowcast roughly halves the error of extrapolation for both cases and deaths, while in steady periods the two are comparable and the added value is small. During fast growth the estimate has an asymmetric downside tail, so decisions between reports should rest on the interval, not the point alone.

## 1 Introduction

Ebola disease is among the most lethal human infections, with case fatality around 50% on average and ranging from roughly 25% to 90% across past outbreaks, varying by virus species and care setting [4, 1, 2]. Bundibugyo virus, the species responsible for the outbreak studied here, has historically been less lethal than Zaire ebolavirus, with case fatality of roughly 25 to 50% in its earlier outbreaks [3, 5]. Ebola outbreaks often occur where surveillance is hardest [6, 7]. Accurate, timely case and death counts are the basic input to any response, yet they are what is hardest to obtain in real time; even the official counts that eventually appear under-ascertain the true burden and are revised as cases are reclassified [8, 9]. The 2026 outbreak studied here, caused by Bundibugyo virus in the Democratic Republic of the Congo and Uganda and declared a public health emergency of international concern in May 2026, is the largest Ebola outbreak yet recorded in the DRC; no licensed vaccine or specific therapeutics exist against Bundibugyo virus [5].

National situation reports describe a period that has closed by the time they are published, and the weekly aggregations built on them can lag by up to a week. Reports are also intermittent, so between them a monitoring function works from the last published figure, which ages until the next one appears. Official reporting is also not always available: in low-capacity, conflict-affected or politically sensitive settings it can be delayed for long periods, patchy, or absent, so a response team may have no recent official figure at all, not merely one that is a few days old. The usual fallback, extending the official curve’s own recent slope, contains no information about what has happened since the last report, and fails at the moments that matter, when an outbreak accelerates or slows.

The approach described here is not specific to Ebola or to this outbreak. In principle it applies wherever case and death figures appear in open reporting, across pathogens and regions, though whether the reporting is dense enough is an empirical question. We validate it on the 2026 DRC Ebola outbreak because it offers a well-reported official series to test against. The question is narrow and testable: does earlier-available open-source reporting improve an estimate of today’s cumulative count over what the official series alone provides?

Real-time forecasting and nowcasting of epidemics is an established field, including model-based forecasts of the West Africa Ebola epidemic [12, 13] and nowcasting methods that correct for reporting delays [10, 11]. A parallel line of work uses digital and open-source signals for earlier outbreak detection, from HealthMap and ProMED [14, 22] to WHO’s Epidemic Intelligence from Open Sources [16] and the EPIWATCH system, and occasionally to estimate epidemiological quantities: Chunara and colleagues estimated reproduction numbers from news and social-media reports that were available up to two weeks earlier than official case data in the 2010 Haitian cholera outbreak [15], though retrospectively. Calibrated, no-look-ahead nowcasts scored against eventual official figures also exist for endemic, data-rich settings, notably the ARGO family for influenza [26]. The present study addresses a different setting: reconstructing and nowcasting reported case and death counts during an acute, emerging outbreak in a low-surveillance environment, where the only official series is a compiled cumulative total.

## 2 Why a nowcast, not a mechanistic model

A compartmental model (SIR, SEIR) or an agent-based model represents the transmission process itself, and is the right tool for estimating a reproduction number (with methods such as EpiEstim or EpiNow2 [24, 25]), testing intervention scenarios, or projecting weeks ahead. Those models also require assumptions about transmission rates, contact structure, susceptibility and reporting, and their medium-term projections are sensitive to those assumptions. Our target is different and narrower: an estimate of the observed cumulative count *as of today*, a few days past the last official figure. Over that short horizon the binding constraint is not knowledge of the transmission mechanism but access to recent data, so a mechanistic model would add parameters and assumptions without adding any observation from the reporting gap. We therefore reconstruct the observed count and project it a short distance, and treat mechanism as complementary rather than absent: the nowcast supplies the calibrated current state that a compartmental or metapopulation model needs as an initial condition, which is the next layer (cross-border importation risk, intervention counterfactuals) rather than part of this estimate.

## 3 Method

This paper describes only the statistical layer that operates on *already-extracted* reported figures; the extraction of structured, source-attributed case and death figures from multilingual open-source text is out of scope here.

Let *O*_*t*_ denote the official cumulative count (cases or deaths) on date *t*. Sources indexed by *s* report values *v*_*s,t*_.

### 1. Source reliability

Sources differ in accuracy, so each is assigned a reliability weight *w*_*s*_ by a source-assessment model that takes no input from the official series, so the weights introduce no dependence on the benchmark the method is scored against. In the backtest these weights are re-estimated at each origin using only reports published on or before that origin, so no component of the evaluation is fit on future data. This weighting is one instance of a broader source-reliability model that is not specific to outbreak reporting and is not detailed here. The validation below does not hinge on it: a generic weighting that simply ranks official-mirror sources above international wire above local reporting reaches the same conclusion, with the more elaborate weighting giving a modest further gain, chiefly on deaths.

### 2. Reconstruction

For each date, the reconstructed value is the reliability-weighted median of the figures reported that day,

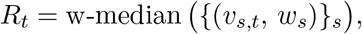

which is robust to any single erroneous report. The series {*R*_*t*_} is then projected onto the monotone non-decreasing cone by isotonic regression [21], since a cumulative count cannot fall. The reconstruction is built only from open, non-official reporting; republications by official agencies are excluded, and are in any case a tiny minority whose removal leaves every result unchanged. Two features of the design make the reconstruction take no input from the compiled official situation report. First, the backtest is strictly no-look-ahead: reports are included only if published on or before the origin date *T* , and each figure is placed at the epidemiological date it refers to, which is on or before its publication date. Second, the compiled official figure for *T* does not appear until several days later, so at *T* it cannot be an input; where open reporting relays official-origin numbers, from a press conference or a provincial team, it conveys them ahead of the compiled situation report, which is the timeliness advantage the method is designed to capture. About half the figures state an explicit observation date; the remainder are assigned their publication date. Restricting the reconstruction to explicitly dated figures alone leaves the direction and significance of the result unchanged; the publication-date fallback is retained because the additional figures improve the estimate, chiefly for deaths. The source list and the extraction method were fixed in advance, and all reporting was collected in real time by the running system; none of the corpus was assembled retrospectively, so the vintage backtest reflects what was genuinely available on each date.

### 3. Nowcast

Let 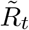be the reconstruction smoothed as the mean of its last three reporting-day values. With *t^*^* the most recent official date and *t*_0_ the present, the nowcast anchors to the most recent official figure and scales it by the reconstruction’s growth over the interval,

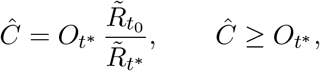

falling back to 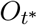unchanged when the reconstruction is unavailable at either endpoint, which does not occur for any origin in this dataset. The *level* is the official figure; the *motion* since then comes from the open-source reconstruction. Because only the ratio 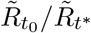 enters, any roughly constant multiplicative offset in the reconstruction cancels: the open-source series tends to trail the official compilation on a rising curve and sits a few per cent below it, but the nowcast takes growth from it, not level, so that offset does not propagate to the estimate. Where no official series is available, the reconstruction itself is the estimate, since it takes no input from the official series.

## 4 Data

The estimand is the national DRC cumulative case and death counts, to match the official series. Uganda reported a small number of linked cases; the validation covers the DRC national series, which accounts for the large majority of the outbreak, and Uganda-specific figures are excluded. Open-source figures are extracted from multilingual reporting on the outbreak. The corpus is 2,205 on-outbreak figures from 67 distinct sources, of which 1,711 are national cumulative counts and 1,591 (case and death counts) enter the reconstruction. Case figures reported as confirmed counts and as total counts are both used; because the nowcast uses only the growth ratio, a roughly constant offset between the two definitions cancels, as above. By tier, aggregator feeds supply 48% of figures and other outlets 31%, with international wire 17% and local reporting 4%; the reliability weighting is what makes this mix usable, since it downweights the low-tier bulk. Official-agency republications are 11 figures (*<* 1%) and are excluded from the reconstruction. The official series is the DRC national situation report (INSP/COUSP), taken from a public mirror and cross-checked against the ECDC outbreak page, which republishes the same national figures. The evaluation window is 23 May to 3 August 2026 for cases (63 official days at or above 100 cases) and 7 June to 3 August for deaths (48 days); the threshold of 100 avoids the unstable percentage errors that small denominators produce early in an outbreak, and the window is frozen at 3 August, the backtest cut-off, with the official series taken at its values as of that freeze. Figure 1 shows the full-sample reconstruction against the official series.

**Figure 1:**
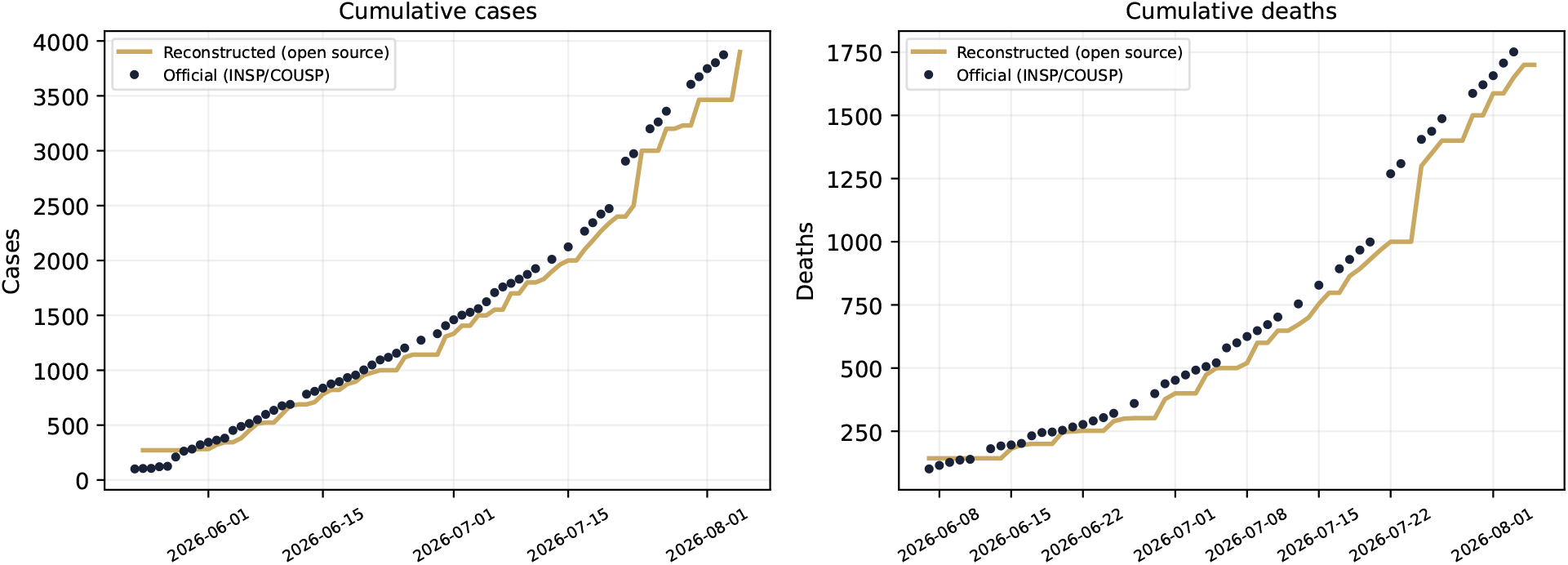
The open-source reconstruction (gold) against the official series (navy points), for cumulative cases and deaths. The reconstruction takes no figures from the compiled official situation report. This panel shows the *full-sample* reconstruction, fitted over all reports, for illustration of how closely open sources can track the curve; the nowcast never uses it. Every quantitative result instead uses the *vintage* reconstruction, rebuilt at each origin date from only the reports published on or before that date, so no future information enters the nowcast.

## 5 Results

We compare three estimates of the current cumulative count in a real-time vintage backtest, using only reports available at each origin (nothing from after it). Each is given the official figure from *L* days earlier and grows it forward across the gap to the origin date; they differ only in the growth rate applied, and each is scored against the figure eventually published for that date. The **official-trend extrapolation** uses the official series’ own recent trend, 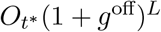, where *g* ^off^ is its compound daily growth over the preceding seven days. The **open-source nowcast** uses the growth of the open-source reconstruction over the interval instead. The **blend** combines the two in log space, log *F* = (1 *− w*) log *F* ^off^ + *w* log *F* ^oss^ with *w* = *L/*(*L* + 3): the inverse-variance posterior mean that stays with the official trend when the gap is short and moves to open-source reporting as it widens [20]. All three are nowcasts of the same quantity; “official-trend” names the one that uses no open-source data, and is itself an extrapolated estimate of today’s figure, not the raw last report. The metric is the median absolute percentage error against the eventual official figure.

The pattern is consistent for both series. When the official figure is fresh (one to three days old) the two are comparable, with simple extrapolation marginally better for cases and no consistent winner for deaths. From four to five days stale for cases, and about six for deaths, the open-source nowcast wins. For cases the improvement is significant from a four-day lag (Wilcoxon signed-rank test [23], *p* = 0.003) and strongly so beyond it; at a one-week lag it cuts median error from 12.6% to 3.9% (*p <* 10^*−*5^). For deaths the gain begins near six days; at a one-week lag it cuts median error from 11.3% to 6.8%, and at eight days from 12.3% to 8.2% (*p* = 0.002). Figure 3 shows the three estimates at 3-, 7- and 10-day lags.

The location of this crossover is the central finding. The method is not intended to improve on a fresh official figure, and it does not: because the benchmark is the eventual official figure itself, a fresh anchor already sits close to the target, leaving little error for any method to reduce. Its value is confined to the interval between the most recent official figure available in real time and the present, and that interval is the normal operating condition. It is set by reporting lag rather than by holes in the record: the compiled official figure for a given day appears only several days later, so at any moment the freshest official count available lags the true one, even for a series that reads as complete once all the reports are in.

### Why the comparison is fair

The official-trend extrapolation and the open-source nowcast use the same minimal projection, an anchor grown by a measured rate, and differ only in the data source for that rate. This is deliberate: holding the model fixed attributes the entire difference to the value of the data, not to model sophistication. The reason a better official-trend model cannot close the gap is informational. No model applied to the headline official series alone, however sophisticated, has a single observation from the interval after the last official report; the open-source reconstruction does. Delay-correction nowcasting [10] recovers information from the reporting triangle of individually dated case reports, but the DRC situation reports publish only compiled cumulative totals, so that machinery has no input here. We checked this directly by replacing the official trend with Holt’s exponential smoothing [19, 18], a standard time-series forecaster, using the damped-trend variant as the conservative choice (an undamped trend overshoots once the cumulative curve decelerates and performs worse). The damped Holt performed on par with simple extrapolation for both series, around 11 to 13% median error at a one-week lag, and neither official-trend forecaster consistently beat the other (simple extrapolation was clearly better than damped Holt for deaths at the longest lags, 13.9% versus 21.9% at ten days). The open-source nowcast beat both from roughly four to five days out for cases and about six days for deaths (Figure 2; Wilcoxon *p ≤* 0.01 at a one-week lag, and smaller beyond). Added model complexity on the official series cannot recover information that is not there. The seven-day window is a standard choice for smoothing a growth-rate estimate over the weekly reporting cycle, and the conclusion does not depend on it. Across windows from three to fourteen days, and under a parameter-free variant that matches the estimation window to the forecast horizon, the crossover is preserved and its direction never reverses: the case crossover falls between three and five days stale and the death crossover between five and eight, with the seven-day default at four and six respectively.

**Figure 2:**
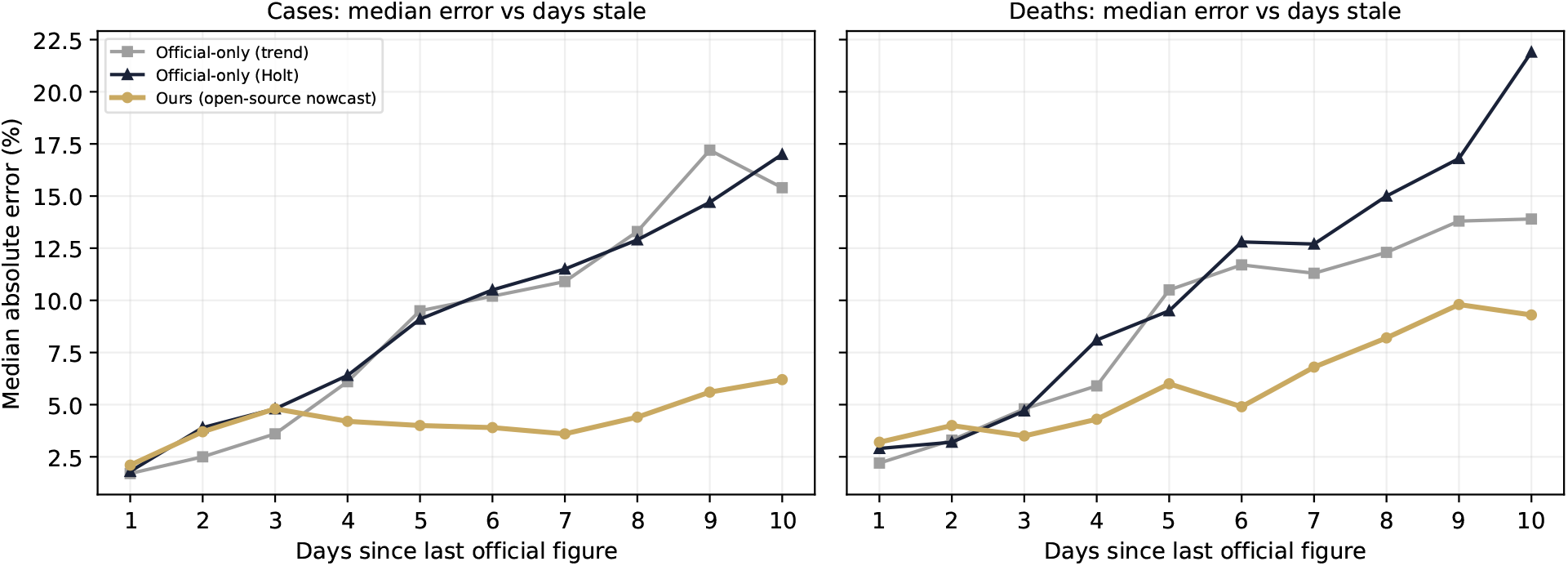
Median absolute percentage error against the eventual official figure, by days since the last official figure, for the two official-trend methods (simple trend and damped Holt) and the open-source nowcast. Both official-trend methods degrade with the lag; the open-source nowcast stays low. Restricted to origins with sufficient official history to fit Holt’s method; Table 1 uses all origins.

**Figure 3:**
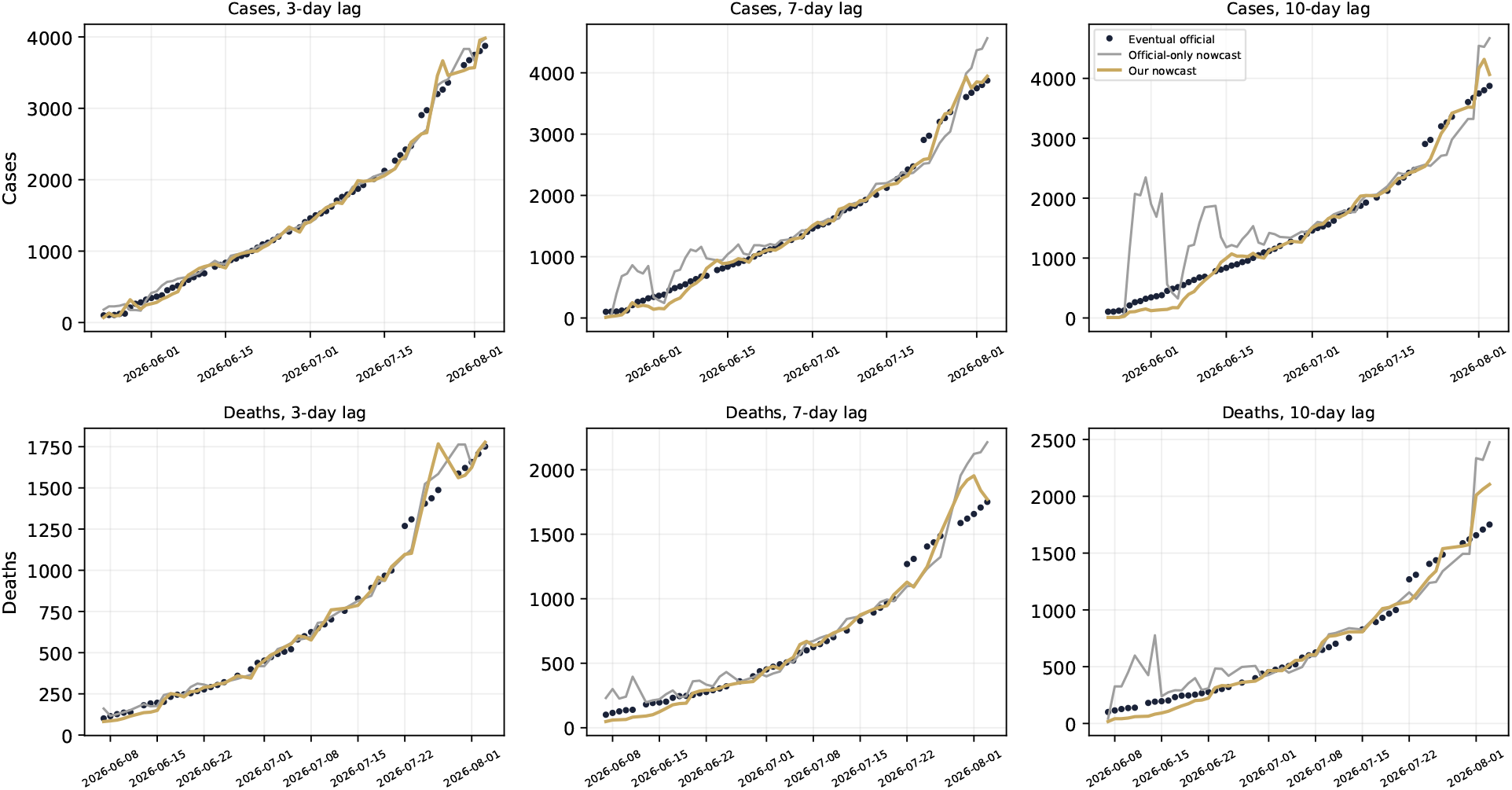
Eventual official figures (navy) against the official-trend extrapolation (grey) and the open-source nowcast (gold), at 3-, 7- and 10-day lags, for cases (top) and deaths (bottom). The open-source nowcast stays close to the eventual figure throughout, while the official-trend extrapolation overshoots during volatile periods. Its large early excursions are the extrapolation compounding the explosive early-phase growth rate, when a small, fast-rising count is projected a week or more forward.

### Where the advantage concentrates: phase transitions

The crossover with the lag is a symptom of a sharper effect. An extrapolation of the official trend can only fail when the trend changes, and it is at those turning points that open-source reports from inside the gap are most informative. Restricting to the established phase, with both the anchor and the target at or above 100 cases to match the scope of the rest of the paper, we stratified every origin by how far the growth rate over the gap departed from the rate the official series was on at the anchor, the departure an extrapolation cannot see, into terciles (Table 2); the stratum label uses the realised movement, while the forecasts remain strictly no-look-ahead. When the epidemic is steady the official extrapolation is accurate and the open-source nowcast adds little. At the turning points the extrapolation is furthest off, at a median error, pooled across origins and horizons (*L* = 1 to 10), of 27.0% for cases and 16.7% for deaths, while the open-source nowcast holds its error far lower, at 13.6% for cases and 7.9% for deaths, roughly half in each case, with the blend close (Table 2). Because the stratifier is the departure of the realised growth from the anchor trend, the top tercile is by construction where extrapolation fails; the informative quantity there is each method’s own error, not the ratio. These medians are descriptive, with no significance test attached to the dependent sample. The advantage is larger still in the explosive early phase, before the count reaches 100, where the official trend can be wrong by well over 100% across a week while the reconstruction observes the surge from inside the gap; those origins sit outside the established-phase scope and are noted separately here to keep the primary result consistent with the rest of the paper. The value is not spread evenly across the outbreak: it is concentrated at the phase transitions, the moments a response most needs a current figure, and is correctly absent when nothing is moving.

**Table 1:**
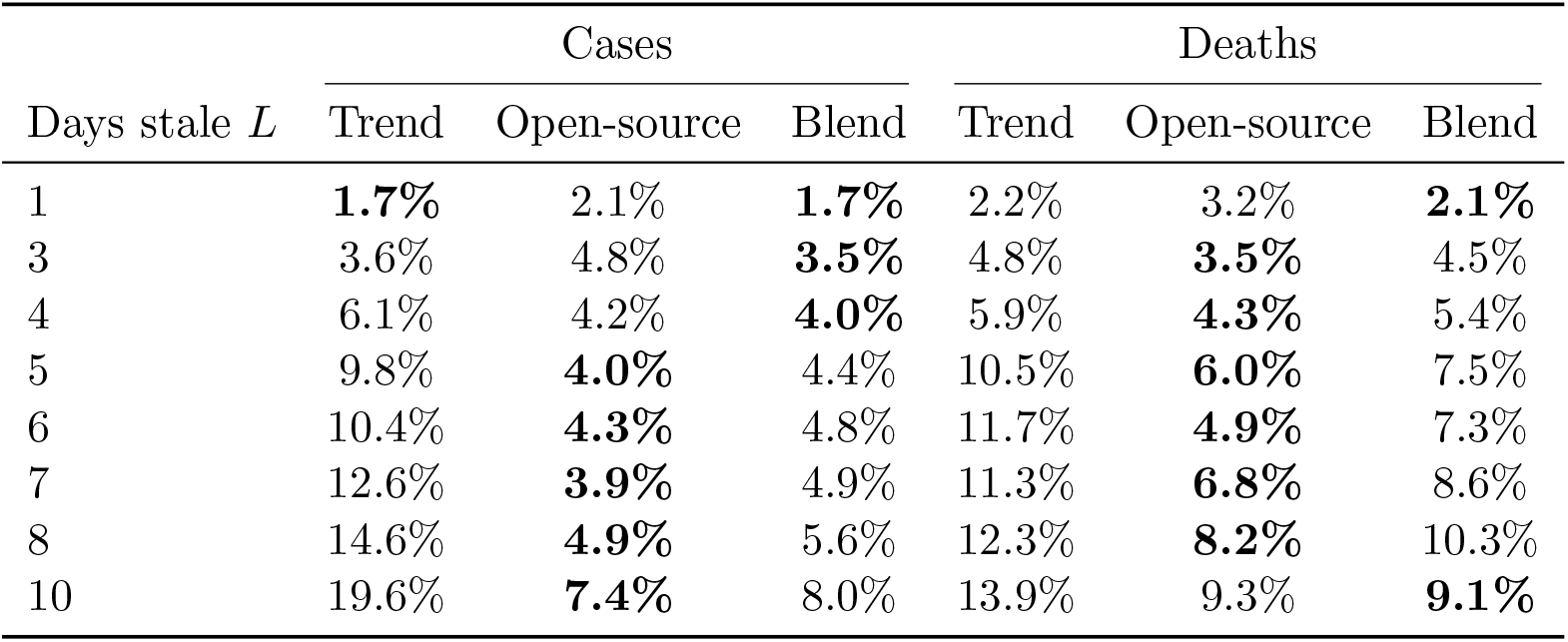
Median absolute percentage error against the eventual official figure, by how many days stale the anchoring official figure is. *N* = 63 origin days for cases (62 at *L* = 10) and 48 for deaths. Bold marks the lower median at each horizon: the official trend when the figure is fresh, the open-source nowcast once it is several days stale, with the blend close to whichever is ahead; where the two are within noise, as for deaths at three to four days stale, the difference is not significant. The open-source nowcast’s advantage over the official trend is significant from four days stale for cases and six for deaths (one-sided paired Wilcoxon signed-rank; values and *p*-values in the text). The official-trend growth rate is estimated over the seven days before the anchor and compounded over the *L*-day gap; official reporting gaps of two to three days make the days-stale label a lower bound on a minority of origins, symmetrically across all methods.

| Days stale $L$ | Cases | | | Deaths | | |
| --- | --- | --- | --- | --- | --- | --- |
|  | Trend | Open-source | Blend | Trend | Open-source | Blend |
| 1 | **1.7%** | 2.1% | **1.7%** | 2.2% | 3.2% | **2.1%** |
| 3 | 3.6% | 4.8% | **3.5%** | 4.8% | **3.5%** | 4.5% |
| 4 | 6.1% | 4.2% | **4.0%** | 5.9% | **4.3%** | 5.4% |
| 5 | 9.8% | **4.0%** | 4.4% | 10.5% | **6.0%** | 7.5% |
| 6 | 10.4% | **4.3%** | 4.8% | 11.7% | **4.9%** | 7.3% |
| 7 | 12.6% | **3.9%** | 4.9% | 11.3% | **6.8%** | 8.6% |
| 8 | 14.6% | **4.9%** | 5.6% | 12.3% | **8.2%** | 10.3% |
| 10 | 19.6% | **7.4%** | 8.0% | 13.9% | 9.3% | **9.1%** |

**Table 2:**
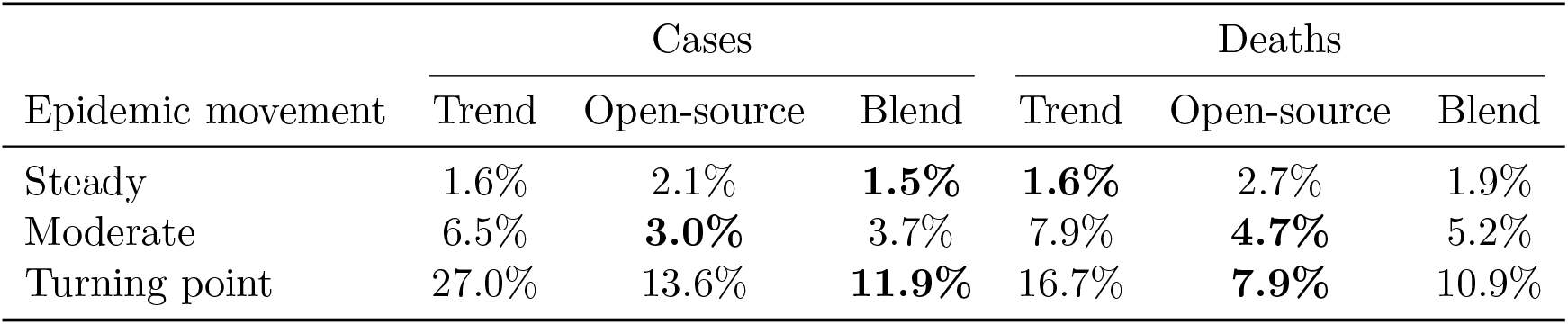
Median absolute percentage error by epidemic movement, for the official trend, the open-source nowcast and their blend, over the established phase (anchor and target both at or above 100 cases). Backtest origins (every origin-day at every horizon *L* = 1 to 10) are pooled and split into terciles by how far the realised growth over the gap departs from the official trend at the anchor; each tercile holds about 190 (origin-day *×* horizon) pairs for cases and 145 for deaths, with each origin day contributing several overlapping pairs. Bold marks the lowest median error in each stratum. The medians are descriptive: because the pairs are dependent and the stratifier is the departure an extrapolation cannot see, no significance test is attached. The forecasts are strictly no-look-ahead.

| Epidemic movement | Cases |  |  | Deaths |  |  |
| --- | --- | --- | --- | --- | --- | --- |
|  | Trend | Open-source | Blend | Trend | Open-source | Blend |
| Steady | 1.6% | 2.1% | **1.5%** | **1.6%** | 2.7% | 1.9% |
| Moderate | 6.5% | **3.0%** | 3.7% | 7.9% | **4.7%** | 5.2% |
| Turning point | 27.0% | 13.6% | **11.9%** | 16.7% | **7.9%** | 10.9% |

### On the blend

The blend defined above, equivalently the classical inverse-mean-squared-error combination of two forecasts [20], follows from a single assumption: an extrapolation’s variance grows with the horizon while the reconstruction’s does not, so its weight *w* = *L/*(*L* + *k*) moves to open-source reporting as the gap widens, and the constant *k* (here 3) is the ratio of the reconstruction’s variance to the extrapolation’s per-day variance. It is never materially worse than the official trend at any lag (Table 1) and is the most accurate at the turning points (Table 2). It is a deployment convenience, not the basis of the finding: the parameter-free open-source nowcast already establishes the result, and the blend’s constant is not tuned to the outcome, since the crossover and the turning-point gain are unchanged for *k* between three and eight and for a variant that sets *w* online from past forecast accuracy.

### Uncertainty of the nowcast

The backtest also yields an empirical error band directly, with no distributional assumption: the quantiles of the nowcast’s signed error at each horizon (Figure 4). The median error is close to zero at every lag, so the nowcast is near-unbiased, and its typical magnitude is the single-digit figures of Table 1. The band is asymmetric and widens with staleness: the upper edge stays within about 10 to 13%, but the lower, under-count tail grows with the lag and is concentrated in the fast-growth origins, where a reconstruction that has not yet caught a surge misses growth that a later official batch reveals. This is the early-phase behaviour noted below, and it is why the interval, rather than the point estimate alone, should inform decisions in the reporting gap: the nowcast errs more often as a slight under-count than as an over-count. The blend narrows this tail materially (Figure 4, lower row). Because the official trend over-counts in the same fast-growth origins where the reconstruction under-counts, combining them roughly halves the eighty per cent interval through the mid-horizons while keeping the median near zero, a further reason to deploy the blend rather than either estimate alone.

**Figure 4:**
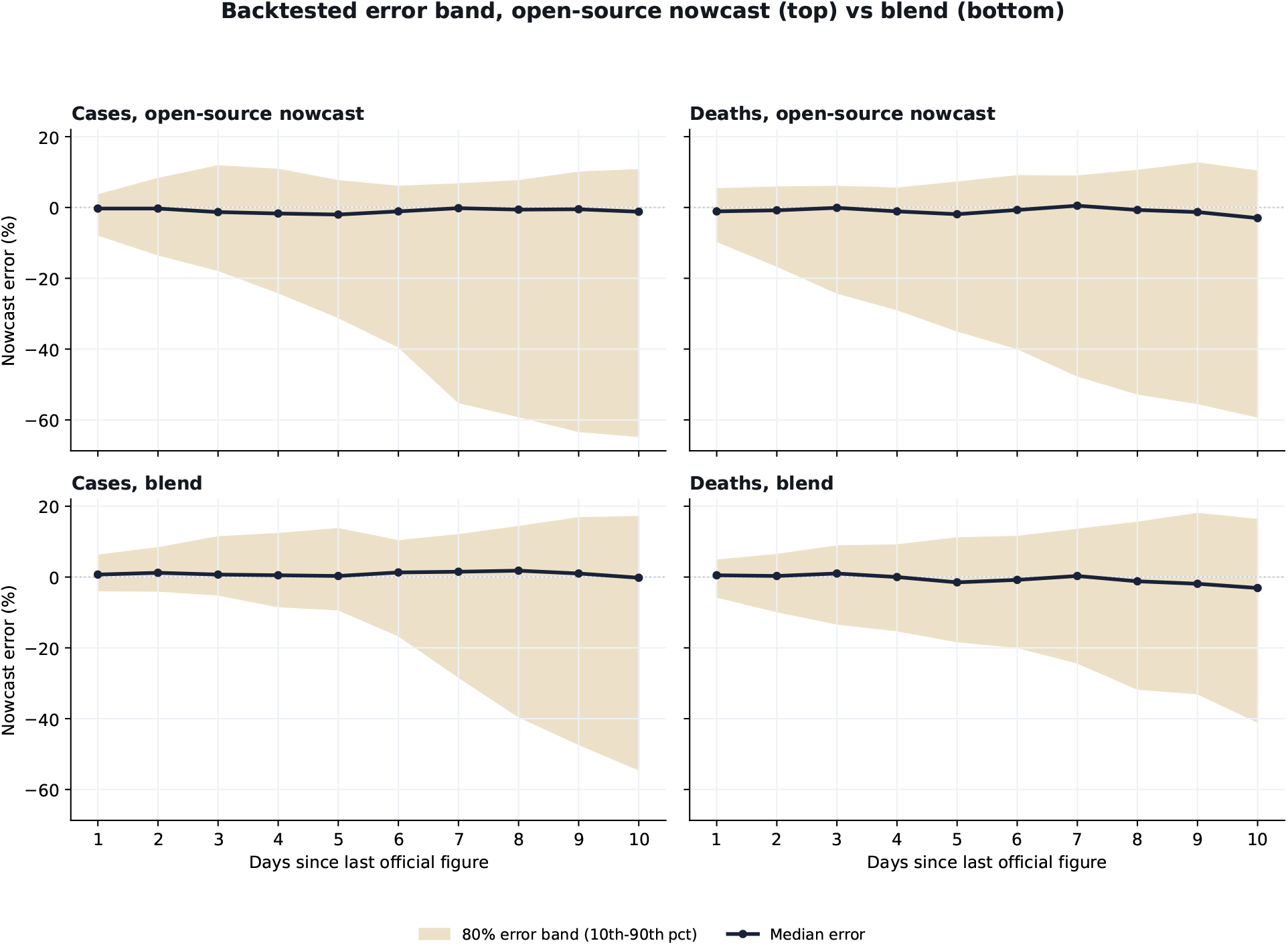
Backtested 80% error band (10th–90th percentile, shaded) and median (line) of the signed error against the eventual official figure, by days since the last official figure, for the open-source nowcast (top row) and the blend (bottom row), over the same origins as Table 1. The median stays near zero at all lags for both. The band is asymmetric, with a downside (under-count) tail that widens with staleness and is concentrated in fast-growth origins; the blend roughly halves that tail through the mid-horizons.

## 6 The operating gap and real-time deployment

The size of the gap the nowcast fills can be read from the source repository’s revision history, in which each situation report is committed on the day it is published. The compiled figure for a given date becomes public a median of two days later. Between reports the last figure is frozen and refreshes a median of once per day (and within two days about 90% of the time), so on a given day the freshest official figure available is a median of two days old over the outbreak, lengthening to about three in the four weeks to 27 August and reaching five at the tail. Each refresh advances the cumulative count by a median of about 4%, and the stale figure a user reads sits a median of about 6% below the count later recorded for that day for cases, and 7 to 8% for deaths, reaching 10 to 17% at the ninetieth percentile. This gap, not the eventual figure, is what a real-time user faces.

Applied at the actual publication lag from 23 May to 24 August, with open-source reports to 27 August, the estimate closes most of it. Against the eventual figure, the stale official figure a user reads is a median 6.3% off for cases and 7.5% for deaths; the combined estimate reduces this to 2.1% and 3.0% and is closer than the stale figure on 95% and 90% of days, and on all eleven evaluated days in the recent three weeks (4 to 24 August), when the lag is longest. A naive extrapolation of the official trend does as well or slightly better over this window (1.9% for cases and 2.3% for deaths), because the deployed weeks were a plateau in which extrapolation is accurate. The improvement is therefore over the unprojected figure a user actually reads, not over a competent extrapolation of it; consistent with the stratified result above, the gain over extrapolation is realised at the transitions, not in the calm.

## 7 Limitations

This validation covers a single outbreak. The series is cumulative and therefore autocorrelated, so the appropriate unit of evidence is the per-horizon sample (63 case-days, 48 death-days), not the pooled count; we therefore do not pool errors across horizons. Even within a horizon consecutive origin days overlap, so the reported *p*-values are anti-conservative; we therefore treat them as descriptive and rest the conclusion on the size and consistency of the median-error gap across horizons and both series. The direction survives two checks against that dependence: a moving-block bootstrap of the per-origin error reductions, with block length equal to the horizon, leaves the case advantage significant from six days stale (*p <* 0.01) and the death advantage significant from six days (*p <* 0.03); and on non-overlapping origins spaced at least a horizon apart the case advantage remains significant from five days (*p <* 0.05, and *p <* 0.02 at all but the nine-day lag), with the death advantage directionally consistent but underpowered on the few spaced points that remain. The comparisons span ten horizons and two series without a formal multiplicity correction, but the conclusion does not rest on any single test: the crossover-adjacent results (cases from four days, *p* = 0.003; deaths from six, *p* = 0.003) survive any standard adjustment, and the finding follows from the consistent direction and magnitude of the gap across horizons. Because both methods are scored against the same eventual figure, their shared denominator does not affect the comparison.

Official counts are themselves imperfect: they under-ascertain the true burden. We benchmark against them because they are the best available reference, not because they are exact, so the method is validated as faster to the official series, not more accurate than it. Two properties of this particular reference bound the claim. It is a single source, the national situation reports, republished unchanged by ECDC, with no independent second series over the study period against which to triangulate, so benchmark uncertainty cannot be estimated from disagreement between sources. And by its published record it is essentially never revised after first publication (one of seventy case-dates, no death-dates), so the eventual figure we score against is also the figure seen in real time; the method is therefore not credited with anticipating later corrections, because there are almost none. Whether the independent open-source reconstruction better approximates the true, under-ascertained burden is an open question we cannot resolve without a gold standard: where the reconstruction diverges from the official series we must score it as error, even though some of that divergence could in principle be closer to the truth.

The estimate applies to the established phase of an outbreak; the explosive early phase and the long tail may behave differently. It is a nowcast of an observed count, not a mechanistic model of transmission. Open-source nowcasting is also prone to overfitting and drift, the failure that undid Google Flu Trends [17]; the safeguard here is that the method is scored by no-look-ahead backtesting against the arriving official figure, not by correlation with a proxy signal.

## 8 Discussion

For a regional preparedness function, the decision-relevant question is where the numbers are now, not where the last report placed them several days ago. A calibrated, independent estimate of the current count, with its error quantified by backtesting, is what the gap between reports leaves missing.

Two points generalise beyond this study, with the caveat that both remain to be tested. First, the estimator is pathogen- and outbreak-agnostic: it requires only that figures appear in open reporting, and nothing in it is specific to Ebola, so it should transfer to other outbreaks, including several monitored concurrently, wherever the reporting is dense enough. Second, official counts are not always available; where they are delayed for long periods or absent, as is common in the settings where outbreaks emerge, the open-source reconstruction becomes the primary quantitative estimate rather than a faster supplement, though where the official pipeline is wholly absent some of the open reporting that relays official-origin figures is absent too. Validating that case is harder because there is no ground truth to test against, so we leave it to future work; here we quantify the gain where an official series exists.

The same open-source data support several extensions: sub-national estimates, imputation for districts that have not yet reported, and calibrated initial conditions for cross-border importation-risk models. The point nowcast is reported here with the empirical error band of Figure 4; a regime-conditional band, tighter in calm periods and wider during acceleration, is a sensible refinement.

## Supporting information

Reproducibility bundle: extracted figures, official series, and backtest code

## Data Availability

All data referred to in the manuscript are openly available. The official DRC national case and death series is public: INRB-UMIE/BDBV2026-Data (https://github.com/INRB-UMIE/BDBV2026-Data), cross-checked against the ECDC outbreak page (https://www.ecdc.europa.eu/en/ebola-outbreak-democratic-republic-congo-and-uganda) and WHO Disease Outbreak News 2026-DON602 (https://www.who.int/emergencies/disease-outbreak-news/item/2026-DON602). The extracted, source-attributed open-source figures and the no-look-ahead backtest code are provided as a supplementary reproducibility bundle with this preprint. An operational implementation runs at https://biosecurity.nymanintelligence.com.

https://www.ecdc.europa.eu/en/ebola-outbreak-democratic-republic-congo-and-uganda

https://www.who.int/emergencies/disease-outbreak-news/item/2026-DON602

## Availability

An operational implementation of the reconstruction and nowcast runs continuously on the outbreak studied here, at biosecurity.nymanintelligence.com.

## Data and code availability

The extracted source-attributed figures, the official series, and the no-look-ahead backtest code are provided as a supplementary reproducibility bundle with this preprint. Two production components are out of scope: the step that produces the figures from source text, and the more detailed source weighting used for the headline results. In their place the bundle uses a simple, fully specified tier weighting (official-mirror sources above international wire above local reporting), and it reproduces the central result and its significance, namely that the open-source nowcast beats the official-trend extrapolation once the official figure is several days stale, with errors slightly larger than the more elaborate weighting, chiefly on deaths.

## Ethics

This study analysed only publicly available, aggregate outbreak counts, taken from official situation reports and public media reporting. No individual-level, patient, or otherwise identifiable data were used and no human participants were involved, so ethics committee approval was not required.

## Author contributions

R.N. is the sole author and performed all aspects of the work.

## Footnotes

A weighted mean does not improve on the weighted median: on the established-phase window its signed bias against the official series is slightly larger (about *−*9% versus *−*7% for cases and *−*9% versus *−*8% for deaths), because the reported figures are skewed toward stale, lower values, which drag the mean down while it also loses the median’s robustness to a single bad report.

The seven-day window matches the weekly reporting cycle and is not a tuned parameter: across windows from three to fourteen days, and under a variant that matches the estimation window to the forecast horizon, the crossover is preserved and its direction never reverses (see “Why the comparison is fair” below).

## References

[1] WHO Ebola Response Team. Ebola Virus Disease in West Africa: the first 9 months of the epidemic and forward projections. New England Journal of Medicine 371(16):1481–1495, 2014. DOI:10.1056/NEJMoa1411100.

[2] Garske T, et al. Heterogeneities in the case fatality ratio in the West African Ebola outbreak 2013–2016. Philosophical Transactions of the Royal Society B 372(1721):20160308, 2017. DOI:10.1098/rstb.2016.0308.

[3] MacNeil A, Farnon EC, Wamala J, et al. Proportion of deaths and clinical features in Bundibugyo Ebola virus infection, Uganda. Emerging Infectious Diseases 16(12):1969–1972, 2010. DOI:10.3201/eid1612.100627.

[4] World Health Organization. Ebola disease (fact sheet, April 2025). who.int/news-room/fact-sheets/detail/ebola-virus-disease (accessed August 2026).

[5] World Health Organization. Ebola disease caused by Bundibugyo virus, Democratic Republic of the Congo and Uganda. Disease Outbreak News 2026-DON602, 16 May 2026. who.int/emergencies/disease-outbreak-news/item/2026-DON602 (accessed August 2026).

[6] Ilunga Kalenga O, Moeti M, Sparrow A, Nguyen VK, Lucey D, Ghebreyesus TA. The ongoing Ebola epidemic in the Democratic Republic of Congo, 2018–2019. New England Journal of Medicine 381(4):373–383, 2019. DOI:10.1056/NEJMsr1904253.

[7] Wells CR, et al. The exacerbation of Ebola outbreaks by conflict in the Democratic Republic of the Congo. PNAS 116(48):24366–24372, 2019. DOI:10.1073/pnas.1913980116.

[8] Tariq A, Roosa K, Mizumoto K, Chowell G. Assessing reporting delays and the effective reproduction number: the Ebola epidemic in DRC, May 2018–January 2019. Epidemics 26:128–133, 2019.

[9] McNamara LA, et al. Ebola surveillance: Guinea, Liberia, and Sierra Leone. MMWR Supplements 65(3):35–43, 2016.

[10] Höhle M, and der Heiden M. Bayesian nowcasting during the STEC O104:H4 outbreak in Germany, 2011. Biometrics 70(4):993–1002, 2014. DOI:10.1111/biom.12194.

[11] McGough SF, Johansson MA, Lipsitch M, Menzies NA. Nowcasting by Bayesian Smoothing: a flexible, generalizable model for real-time epidemic tracking. PLOS Computational Biology 16(4):e1007735, 2020. DOI:10.1371/journal.pcbi.1007735.

[12] Chowell G, et al. Perspectives on model forecasts of the 2014–2015 Ebola epidemic in West Africa: lessons and the way forward. BMC Medicine 15:42, 2017. DOI:10.1186/s12916-017-0811-y.

[13] Funk S, Camacho A, Kucharski AJ, Lowe R, Eggo RM, Edmunds WJ. Assessing the performance of real-time epidemic forecasts: a case study of Ebola in the Western Area Region of Sierra Leone, 2014–15. PLOS Computational Biology 15(2):e1006785, 2019. DOI:10.1371/journal.pcbi.1006785.

[14] Brownstein JS, Freifeld CC, Reis BY, Mandl KD. Surveillance sans frontières: internet-based emerging infectious disease intelligence and the HealthMap project. PLoS Medicine 5(7):e151, 2008. DOI:10.1371/journal.pmed.0050151.

[15] Chunara R, Andrews JR, Brownstein JS. Social and news media enable estimation of epidemiological patterns early in the 2010 Haitian cholera outbreak. American Journal of Tropical Medicine and Hygiene 86(1):39–45, 2012. DOI:10.4269/ajtmh.2012.11-0597.

[16] Williams GS, Koua EL, Abdelmalik P, et al. Evaluation of the Epidemic Intelligence from Open Sources (EIOS) system for the early detection of outbreaks and health emergencies in the African region. BMC Public Health 25:857, 2025. DOI:10.1186/s12889-025-21998-9.

[17] Lazer D, Kennedy R, King G, Vespignani A. The parable of Google Flu: traps in big data analysis. Science 343(6176):1203–1205, 2014. DOI:10.1126/science.1248506.

[18] Hyndman RJ, Athanasopoulos G. Forecasting: Principles and Practice, 3rd edition. OTexts, 2021.

[19] Holt CC. Forecasting seasonals and trends by exponentially weighted moving averages. International Journal of Forecasting 20(1):5–10, 2004. DOI:10.1016/j.ijforecast.2003.09.015.

[20] Bates JM, Granger CWJ. The combination of forecasts. Operational Research Quarterly 20(4):451–468, 1969.

[21] Barlow RE, Bartholomew DJ, Bremner JM, Brunk HD. Statistical Inference Under Order Restrictions: The Theory and Application of Isotonic Regression. Wiley, 1972.

[22] Madoff LC. ProMED-mail: an early warning system for emerging diseases. Clinical Infectious Diseases 39(2):227–232, 2004.

[23] Wilcoxon F. Individual comparisons by ranking methods. Biometrics Bulletin 1(6):80–83, 1945.

[24] Cori A, Ferguson NM, Fraser C, Cauchemez S. A new framework and software to estimate time-varying reproduction numbers during epidemics. American Journal of Epidemiology 178(9):1505–1512, 2013. DOI:10.1093/aje/kwt133.

[25] Abbott S, Hellewell J, Thompson RN, et al. Estimating the time-varying reproduction number of SARS-CoV-2 using national and subnational case counts. Wellcome Open Research 5:112, 2020. DOI:10.12688/wellcomeopenres.16006.2.

[26] Yang S, Santillana M, Kou SC. Accurate estimation of influenza epidemics using Google search data via ARGO. Proceedings of the National Academy of Sciences 112(47):14473–14478, 2015. DOI:10.1073/pnas.1515373112.

