## Supplementary material for "Nowcasting outbreak case and death counts from open-source reporting A validation on the 2026 Ebola (Bundibugyo) outbreak in the Democratic Republic of the Congo": Reproducibility bundle: extracted figures, official series, and backtest code: benchmark.py

```
"""Backtest: open-source nowcast vs projecting the last official figure forward.

No look-ahead. At each origin the official figure from L days earlier is given to both
methods, which estimate the value L days later (the eventual official figure):
  official-only : anchor * (1 + recent 7-day official growth)^L
  ours          : anchor * (reconstruction growth over the interval)
Median absolute % error by horizon, one-sided paired Wilcoxon test.

Uses the simple tier reconstruction here; the paper's headline numbers use the
production source weighting and are slightly better. Same conclusion either way.

Run:  python3 benchmark.py
"""
import numpy as np
from datetime import date, timedelta
from reconcile import load
from validate import load_official
from nowcast import vintage_curve, value_asof, recent_growth
from scipy.stats import wilcoxon

GROUPS = {"cases": {"confirmed_cases", "total_cases"}, "deaths": {"deaths"}}
OFFICIAL = {"cases": ("official_cases.csv", "national_cumulative_confirmed_cases"),
            "deaths": ("official_deaths.csv", "national_cumulative_confirmed_deaths")}


def dd(s): return date(*map(int, s.split("-")))
def shift(s, n): return (dd(s) - timedelta(days=n)).isoformat()
def med(xs): return float(np.median(xs)) if xs else None
def ape(est, o): return abs(est - o) / o


def smooth_value(curve, d, k=3):
    ks = [x for x in sorted(curve) if x <= d]
    return sum(curve[x] for x in ks[-k:]) / len(ks[-k:]) if ks else None


def main():
    figs = load()
    for metric in ("cases", "deaths"):
        official = load_official(*OFFICIAL[metric]); spec = GROUPS[metric]
        dates = [d for d in sorted(official) if official[d] >= 100]
        vint = {d: vintage_curve(figs, spec, d) for d in dates}   # tier reconstruction
        print(f"\n===== {metric.upper()}  ({dates[0]} to {dates[-1]}, N={len(dates)}) =====")
        print(f"{'L':>3} | {'official-only':>13} {'ours':>7} | {'ours<off p':>10}")
        for L in range(1, 11):
            eo, ea = [], []
            for D in dates:
                A = shift(D, L); base = value_asof(official, A); truth = official.get(D)
                if not base or not truth:
                    continue
                g = recent_growth(official, A, w=7)
                rd, ra = smooth_value(vint[D], D), smooth_value(vint[D], A)
                ours = base * (rd / ra) if (rd and ra) else base
                eo.append(ape(base * ((1 + g) ** L), truth)); ea.append(ape(ours, truth))
            p = wilcoxon(eo, ea, alternative="greater").pvalue if len(eo) >= 6 else float("nan")
            print(f"{L:>3} | {100*med(eo):12.1f}% {100*med(ea):6.1f}% | {p:>10.4f}")


if __name__ == "__main__":
    main()
```
