## Supplementary material for "Nowcasting outbreak case and death counts from open-source reporting A validation on the 2026 Ebola (Bundibugyo) outbreak in the Democratic Republic of the Congo": Reproducibility bundle: extracted figures, official series, and backtest code: deployment.py

```
"""Reporting lag and real-time deployment, on data to late August 2026.

(A) Reporting lag of the official INSP national series, from publication dates
    (insp_publication_dates.json: each observation date mapped to the day the
    situation report carrying it was published, taken from the source repository's
    commit history). How old the freshest available figure is on a given day, how
    often it refreshes, and how far the stale figure a user reads sits below the
    count later recorded for that day.

(B) Real-time deployment: at the ACTUAL publication lag, compare the stale official
    figure, a naive official-trend extrapolation, the open-source nowcast and a simple
    blend against the eventual official figure.

Uses the live snapshots (official to 24 Aug, open-source figures to 27 Aug) and the
tier reconstruction; the paper's exact numbers use the production source weighting.

Run:  python3 deployment.py
"""
import os, json, math
import numpy as np
from datetime import date, timedelta
from collections import Counter

import reconcile
reconcile.IN = os.path.join(os.path.dirname(__file__), "extracted_epi_figures_live.json")
from reconcile import load
from validate import load_official
from nowcast import vintage_curve, value_asof, recent_growth

HERE = os.path.dirname(__file__)
PUB = json.load(open(os.path.join(HERE, "insp_publication_dates.json")))
PUBDATES = {k: date(*map(int, v.split("-"))) for k, v in PUB.items()}
OFFICIAL = {"cases": ("official_cases_live.csv", "national_cumulative_confirmed_cases"),
            "deaths": ("official_deaths_live.csv", "national_cumulative_confirmed_deaths")}
GROUPS = {"cases": {"confirmed_cases", "total_cases"}, "deaths": {"deaths"}}
FROZEN_END = date(2026, 8, 3)
END = date(2026, 8, 24)


def dd(s): return date(*map(int, s.split("-")))
def med(xs): return float(np.median(xs)) if xs else None
def ape(e, o): return abs(e - o) / o
def smooth_value(curve, d, k=3):
    ks = [x for x in sorted(curve) if x <= d]
    return sum(curve[x] for x in ks[-k:]) / len(ks[-k:]) if ks else None


def reporting_lag(metric):
    official = load_official(*OFFICIAL[metric])
    obs = sorted(o for o in official if o in PUBDATES and official[o] >= 100)
    lag = {o: (PUBDATES[o] - dd(o)).days for o in obs}
    early = [v for o, v in lag.items() if dd(o) <= FROZEN_END]
    recent = [v for o, v in lag.items() if dd(o) > FROZEN_END]
    start = min(PUBDATES[o] for o in obs)
    fresh = []
    T = start
    while T <= date(2026, 8, 27):
        avail = [o for o in obs if PUBDATES[o] <= T and dd(o) <= T]
        fresh.append((T, max(avail, key=dd) if avail else None))
        T += timedelta(days=1)
    stale_age = [(T - dd(fr)).days for T, fr in fresh if fr]
    changes = [i for i in range(1, len(fresh)) if fresh[i][1] != fresh[i-1][1]]
    gaps = [changes[i]-changes[i-1] for i in range(1, len(changes))]
    seq = [o for o in obs]
    jumps = [100*(official[seq[i]]-official[seq[i-1]])/official[seq[i-1]] for i in range(1, len(seq))]
    behind = []
    for T, fr in fresh:
        if not fr:
            continue
        cur = [official[o] for o in seq if dd(o) <= T]
        if cur and official[fr] > 0:
            behind.append(100*(cur[-1]-official[fr])/official[fr])
    print(f"  publication lag (obs date -> published): median {med(early):.0f}d through 3 Aug, "
          f"{med(recent):.0f}d recently")
    print(f"  freshest figure age on a given day: median {med(stale_age):.0f}d, max {max(stale_age)}d")
    print(f"  refresh interval dist (days): {dict(sorted(Counter(gaps).items()))}")
    print(f"  size of each refresh: median {med(jumps):.1f}%")
    print(f"  stale figure sits below current count: median {med(behind):.1f}%, 90th pct {np.percentile(behind,90):.1f}%")


def deployment(metric, figs):
    official = load_official(*OFFICIAL[metric]); spec = GROUPS[metric]
    alldates = sorted(official)
    dates = [d for d in alldates if official[d] >= 100 and dd(d) <= END]
    vint = {d: vintage_curve(figs, spec, d) for d in dates}

    def block(dsub, label):
        rows = []
        for D in dsub:
            Dt = dd(D)
            avail = [d2 for d2 in alldates if dd(d2) < Dt and d2 in PUBDATES and PUBDATES[d2] <= Dt]
            if not avail:
                continue
            anchor = max(avail, key=dd); L = (Dt - dd(anchor)).days
            base = official[anchor]; truth = official[D]
            g = recent_growth(official, anchor, w=7); offtrend = base*((1+g)**L)
            rd, ra = smooth_value(vint[D], D), smooth_value(vint[D], anchor)
            osint = base*(rd/ra) if (rd and ra) else offtrend
            if rd and ra:
                w = L/(L+3); blend = base*math.exp((1-w)*math.log((1+g)**L)+w*math.log(rd/ra))
            else:
                blend = offtrend
            rows.append((L, ape(base, truth), ape(offtrend, truth), ape(osint, truth), ape(blend, truth)))
        n = len(rows)
        print(f"  {label}: n={n}, median real lag {med([r[0] for r in rows]):.0f}d")
        print(f"    stale {100*med([r[1] for r in rows]):.1f}% | off-trend {100*med([r[2] for r in rows]):.1f}% | "
              f"nowcast {100*med([r[3] for r in rows]):.1f}% | blend {100*med([r[4] for r in rows]):.1f}%")
        print(f"    blend closer than stale figure on {100*sum(1 for r in rows if r[4]<r[1])/n:.0f}% of days")
    block(dates, "23 May-24 Aug")
    block([d for d in dates if dd(d) > FROZEN_END], "4-24 Aug (recent)")


def main():
    figs = load()
    for metric in ("cases", "deaths"):
        print(f"\n===== {metric.upper()} =====")
        reporting_lag(metric)
        deployment(metric, figs)


if __name__ == "__main__":
    main()
```
