## Supplementary material for "Nowcasting outbreak case and death counts from open-source reporting A validation on the 2026 Ebola (Bundibugyo) outbreak in the Democratic Republic of the Congo": Reproducibility bundle: extracted figures, official series, and backtest code: LICENSE.txt

Reproduction bundle for "Nowcasting outbreak case and death counts from open-source
reporting: a validation on the 2026 Ebola (Bundibugyo) outbreak in the Democratic
Republic of the Congo" (Nyman, 2026).

Copyright (c) 2026 Rickard Nyman / Nyman Intelligence.

Code (all *.py files): MIT License.

  Permission is hereby granted, free of charge, to any person obtaining a copy of
  this software and associated files, to deal in the software without restriction,
  including the rights to use, copy, modify, merge, publish, and distribute it,
  subject to the inclusion of the above copyright notice. THE SOFTWARE IS PROVIDED
  "AS IS", WITHOUT WARRANTY OF ANY KIND.

Data (extracted_epi_figures.json, official_cases.csv, official_deaths.csv):
Creative Commons Attribution 4.0 International (CC BY 4.0).
You are free to share and adapt the data with attribution to the author.

The production extraction and source-weighting components are not included and are
not licensed here; they remain proprietary to Nyman Intelligence. This bundle uses a
simple tier weighting in their place.
