## Supplementary material for "Nowcasting outbreak case and death counts from open-source reporting A validation on the 2026 Ebola (Bundibugyo) outbreak in the Democratic Republic of the Congo": Reproducibility bundle: extracted figures, official series, and backtest code: nowcast.py

```
"""Backtested nowcast: can a real-time open-source reconstruction (using only reports
available up to day D) estimate the current cumulative earlier than, and about as
accurately as, the official series once its own reporting lag is corrected?

  1. Vintages: for each day D, reconstruct using only figures published on or before D;
     R(D) is the real-time edge value.
  2. Lag: on a training window, find the shift tau for which official(D-tau) best
     matches R(D).
  3. Nowcast: project R(D) forward tau days at the recent growth rate; credible
     interval from training residuals.
  4. Score R(D) and N(D) against official(D), and measure lead time.

Run:  python3 nowcast.py
"""
import os, json, statistics as st
from datetime import datetime, timedelta
from reconcile import load, reconcile_metric
from validate import load_official

HERE = os.path.dirname(__file__)
GROUPS = {"cases": {"confirmed_cases", "total_cases"}, "deaths": {"deaths"}}
OFFICIAL = {"cases": ("official_cases.csv", "national_cumulative_confirmed_cases"),
            "deaths": ("official_deaths.csv", "national_cumulative_confirmed_deaths")}


def dd(s):
    return datetime.strptime(s, "%Y-%m-%d").date()


def daterange(a, b):
    d = dd(a)
    while d <= dd(b):
        yield d.isoformat()
        d += timedelta(days=1)


def vintage_curve(figs, spec, asof):
    """Reconciled cumulative dict {date: value} using only figures published <= asof."""
    sub = [f for f in figs if f.get("_pubdate") and f["_pubdate"] <= asof]
    c = reconcile_metric(sub, spec)
    return {d: v for d, v, *_ in c}


def value_asof(curve, d):
    ks = [k for k in curve if k <= d]
    return curve[max(ks)] if ks else None


def recent_growth(curve, d, w=7):
    """Daily multiplicative growth of the reconstructed cumulative over last w days up to d."""
    ks = sorted(k for k in curve if k <= d)
    if len(ks) < 2:
        return 0.0
    hi = curve[ks[-1]]
    past = [k for k in ks if k <= (dd(d) - timedelta(days=w)).isoformat()]
    lo = curve[past[-1]] if past else curve[ks[0]]
    span = max(1, (dd(ks[-1]) - dd(past[-1] if past else ks[0])).days)
    if lo <= 0 or hi <= 0:
        return 0.0
    return (hi / lo) ** (1.0 / span) - 1.0


def estimate_lag(vint, official, dates, max_tau=10):
    """Median tau such that official(D - tau) ~= R(D) across training dates."""
    taus = []
    for D in dates:
        R = vint.get(D)
        if not R:
            continue
        best, bt = 1e9, 0
        for tau in range(0, max_tau + 1):
            Dm = (dd(D) - timedelta(days=tau)).isoformat()
            o = value_asof(official, Dm)
            if o and abs(o - R) / R < best:
                best, bt = abs(o - R) / R, tau
        taus.append(bt)
    return st.median(taus) if taus else 0


def backtest(label):
    figs = load()
    spec = GROUPS[label]
    offf, offc = OFFICIAL[label]
    official = load_official(offf, offc)

    # backtest window: established phase only (official >= 100), daily
    start = min((d for d in sorted(official) if official[d] >= 100), default=None)
    end = max(official)
    if not start:
        print(f"{label}: no established phase"); return None
    dates = [d for d in daterange(start, end)]

    # build a full point-in-time vintage curve per eval date (reports published <= D)
    print(f"[{label}] building {len(dates)} daily vintages ...")
    vint_curve = {D: vintage_curve(figs, spec, D) for D in dates}
    vint = {D: value_asof(vint_curve[D], D) for D in dates}        # real-time edge value R(D)
    full = vintage_curve(figs, spec, end)

    # split train/test by time (first 60% train)
    cut = dates[int(len(dates) * 0.6)]
    train = [d for d in dates if d <= cut]
    test = [d for d in dates if d > cut]
    tau = estimate_lag(vint, official, train)
    tau_i = int(round(tau))
    print(f"[{label}] estimated reconstruction lag tau = {tau} days (trained on {len(train)} days)")

    def nowcast(D):
        """Pure-OSINT nowcast: project the real-time edge forward by tau (no official input)."""
        R = vint.get(D)
        if not R:
            return None
        g = recent_growth(full, D)
        return R * ((1.0 + g) ** tau_i)

    def anchored(D, L):
        """Anchored nowcast: official(D-L) scaled by OSINT growth over the unpublished tail (D-L, D].
        Uses only official data available L days ago plus the real-time OSINT vintage as of D."""
        anchor = (dd(D) - timedelta(days=L)).isoformat()
        base = value_asof(official, anchor)
        rd, ra = vint.get(D), value_asof(vint_curve[D], anchor)
        if not base or not rd or not ra:
            return None
        return base * (rd / ra)

    # training residuals for a credible interval on the nowcast
    tr = [(nowcast(D) - official[D]) / official[D]
          for D in train if vint.get(D) and D in official]
    lo_q = sorted(tr)[max(0, int(0.1 * len(tr)) - 1)] if tr else 0
    hi_q = sorted(tr)[min(len(tr) - 1, int(0.9 * len(tr)))] if tr else 0

    def score(fn, ds):
        errs = []
        for D in ds:
            o = official.get(D)
            if not o:
                o = value_asof(official, D)
            est = fn(D)
            if est and o:
                errs.append((est - o) / o)
        ape = sorted(abs(e) for e in errs)
        bias = sorted(errs)
        n = len(errs)
        return n, 100 * ape[n // 2], 100 * bias[n // 2]

    print(f"\n[{label}] TEST-period accuracy vs official (n days held out):")
    methods = [("raw real-time reconstruction", lambda D: vint.get(D)),
               ("pure-OSINT nowcast (no official)", nowcast),
               ("ANCHORED nowcast, official lag L=3", lambda D: anchored(D, 3)),
               ("ANCHORED nowcast, official lag L=7", lambda D: anchored(D, 7))]
    now_mape = None
    for name, fn in methods:
        n, mape, bias = score(fn, test)
        if name.startswith("pure-OSINT"):
            now_mape = mape
        print(f"   {name:36s} n={n:3d}  median APE {mape:5.1f}%   median bias {bias:+5.1f}%")
    print(f"   pure-OSINT nowcast 80% credible band (training): {100*lo_q:+.0f}% .. {100*hi_q:+.0f}%")

    # lead time: first day each series crosses a level
    print(f"\n[{label}] lead time (days OSINT nowcast crosses a level BEFORE official):")
    leads = []
    levels = [v for v in (200, 500, 1000, 1500, 2000, 3000) if v <= official[end]]
    for V in levels:
        def first_cross(getter, ds):
            for D in ds:
                val = getter(D)
                if val and val >= V:
                    return D
            return None
        d_now = first_cross(nowcast, dates)
        d_off = next((d for d in sorted(official) if official[d] >= V), None)
        if d_now and d_off:
            lead = (dd(d_off) - dd(d_now)).days
            leads.append(lead)
            print(f"   {V:5d}: nowcast {d_now}   official {d_off}   lead {lead:+d} d")
    if leads:
        print(f"   median lead: {st.median(leads):+.1f} days")

    return {"label": label, "tau": tau_i, "dates": dates, "test_start": test[0] if test else None,
            "raw": vint, "nowcast": {D: nowcast(D) for D in dates},
            "anchored7": {D: anchored(D, 7) for D in dates}, "official": official,
            "band": (lo_q, hi_q), "test_mape": now_mape}


def plot(results):
    import matplotlib
    matplotlib.use("Agg")
    import matplotlib.pyplot as plt
    import matplotlib.dates as mdates
    labels = [l for l in ("cases", "deaths") if results.get(l)]
    fig, axes = plt.subplots(1, len(labels), figsize=(6.6 * len(labels), 5.2), squeeze=False)
    for ax, label in zip(axes[0], labels):
        r = results[label]
        ds = [d for d in r["dates"]]
        x = [dd(d) for d in ds]
        off = [value_asof(r["official"], d) for d in ds]
        ax.plot(x, off, color="#c0392b", lw=2.4, ls=(0, (5, 2)), label="Official (eventual truth)", zorder=5)
        ax.plot(x, [r["raw"].get(d) for d in ds], color="#9aa3b0", lw=1.6, label="Raw real-time OSINT", zorder=3)
        ax.plot(x, [r["nowcast"].get(d) for d in ds], color="#1f2430", lw=2.2, label="Pure-OSINT nowcast", zorder=6)
        ax.plot(x, [r["anchored7"].get(d) for d in ds], color="#2e86c1", lw=2.0, label="Anchored nowcast (L=7)", zorder=6)
        if r["test_start"]:
            ax.axvline(dd(r["test_start"]), color="#c8ccd4", lw=1, ls=":")
            ax.text(dd(r["test_start"]), ax.get_ylim()[1] * 0.02, " held-out test", fontsize=7.5, color="#8a93a4")
        ax.set_title(label.capitalize(), fontsize=13, fontweight="bold", loc="left")
        ax.set_ylabel("Cumulative"); ax.xaxis.set_major_formatter(mdates.DateFormatter("%d %b"))
        ax.grid(True, color="#eef1f5"); ax.tick_params(labelsize=8.5)
        for s in ("top", "right"):
            ax.spines[s].set_visible(False)
    h, l = axes[0][0].get_legend_handles_labels()
    fig.legend(h, l, loc="lower center", ncol=4, frameon=False, fontsize=9, bbox_to_anchor=(0.5, -0.02))
    fig.suptitle("Backtested nowcast: real-time estimate vs official (never seen by the model)",
                 fontsize=14, fontweight="bold", y=1.0)
    fig.tight_layout(rect=[0, 0.05, 1, 0.95])
    out = os.path.join(HERE, "nowcast_backtest.png")
    fig.savefig(out, dpi=150, bbox_inches="tight", facecolor="white")
    print(f"wrote {out}")


def main():
    results, summ = {}, {}
    for label in ("cases", "deaths"):
        r = backtest(label)
        if r:
            results[label] = r
            summ[label] = {"tau": r["tau"], "band": r["band"], "test_mape": r["test_mape"]}
        print()
    if results:
        try:
            plot(results)
        except ImportError:
            print("(matplotlib not installed; skipping the plot)")
    json.dump(summ, open(os.path.join(HERE, "nowcast_summary.json"), "w"), indent=1)


if __name__ == "__main__":
    main()
```
