## Supplementary material for "Nowcasting outbreak case and death counts from open-source reporting A validation on the 2026 Ebola (Bundibugyo) outbreak in the Democratic Republic of the Congo": Reproducibility bundle: extracted figures, official series, and backtest code: phase_transition.py

```
"""Where does the open-source nowcast add value? Performance by epidemic MOVEMENT.

At a phase transition the official series' recent trend (what an extrapolation projects)
diverges from what actually happens over the gap. That is where extrapolation must fail
and where open-source reports from inside the gap help most.

For every backtest origin (D, L) we measure the SURPRISE: the gap between the daily
growth rate the official series was on at the anchor (g_before, trailing 7 days, what an
extrapolation projects) and the daily growth rate that actually occurred over the L-day
gap (g_gap). Origins are split into terciles of |surprise| and we compare median absolute
% error for official-only extrapolation, the open-source nowcast, and a simple blend of
the two. The stratum label uses the realised movement (hindsight); the forecasts are
strictly no-look-ahead.

Tier reconstruction, as in benchmark.py; the paper's exact numbers use the production
source weighting.

Run:  python3 phase_transition.py
"""
import math
import numpy as np
from datetime import date, timedelta
from reconcile import load
from validate import load_official
from nowcast import vintage_curve, value_asof, recent_growth
from scipy.stats import wilcoxon

GROUPS = {"cases": {"confirmed_cases", "total_cases"}, "deaths": {"deaths"}}
OFFICIAL = {"cases": ("official_cases.csv", "national_cumulative_confirmed_cases"),
            "deaths": ("official_deaths.csv", "national_cumulative_confirmed_deaths")}


def dd(s): return date(*map(int, s.split("-")))
def shift(s, n): return (dd(s) - timedelta(days=n)).isoformat()
def med(xs): return float(np.median(xs)) if xs else None
def ape(e, o): return abs(e - o) / o
def smooth_value(curve, d, k=3):
    ks = [x for x in sorted(curve) if x <= d]
    return sum(curve[x] for x in ks[-k:]) / len(ks[-k:]) if ks else None


def main():
    figs = load()
    for metric in ("cases", "deaths"):
        official = load_official(*OFFICIAL[metric]); spec = GROUPS[metric]
        dates = [d for d in sorted(official) if official[d] >= 100]
        vint = {d: vintage_curve(figs, spec, d) for d in dates}
        rows = []
        for L in range(1, 11):
            for D in dates:
                A = shift(D, L); base = value_asof(official, A); truth = official.get(D)
                if not base or not truth or base < 100:   # established phase at both anchor and target
                    continue
                g_before = recent_growth(official, A, w=7)
                g_gap = (truth / base) ** (1.0 / L) - 1.0
                offtrend = base * ((1 + g_before) ** L)
                rd, ra = smooth_value(vint[D], D), smooth_value(vint[D], A)
                osint = base * (rd / ra) if (rd and ra) else base
                if rd and ra:
                    w = L / (L + 3)
                    blend = base * math.exp((1 - w) * math.log((1 + g_before) ** L) + w * math.log(rd / ra))
                else:
                    blend = offtrend
                rows.append({"surprise": abs(g_gap - g_before), "off": ape(offtrend, truth),
                             "oss": ape(osint, truth), "bl": ape(blend, truth)})
        s = sorted(rows, key=lambda r: r["surprise"]); n = len(s)
        strata = [("steady", s[:n//3]), ("moderate", s[n//3:2*n//3]), ("turning point", s[2*n//3:])]
        print(f"\n===== {metric.upper()}: median APE by epidemic movement =====")
        print(f"  {'stratum':<14} {'n':>4} {'off-trend':>10} {'nowcast':>8} {'blend':>7}  {'p(oss<off)':>11}")
        for name, grp in strata:
            off = [r["off"] for r in grp]; oss = [r["oss"] for r in grp]; bl = [r["bl"] for r in grp]
            try:
                p = wilcoxon(off, oss, alternative="greater").pvalue if any(a != b for a, b in zip(off, oss)) else float("nan")
            except ValueError:
                p = float("nan")
            print(f"  {name:<14} {len(grp):>4} {100*med(off):>9.1f}% {100*med(oss):>7.1f}% {100*med(bl):>6.1f}%  {p:>11.4f}")


if __name__ == "__main__":
    main()
```
