## Supplementary material for "Nowcasting outbreak case and death counts from open-source reporting A validation on the 2026 Ebola (Bundibugyo) outbreak in the Democratic Republic of the Congo": Reproducibility bundle: extracted figures, official series, and backtest code: README.md

# Reproduction bundle: open-source nowcasting vs official figures

This bundle reproduces the central result of the paper "Nowcasting outbreak case and
death counts from open-source reporting: a validation on the 2026 Ebola (Bundibugyo)
outbreak in the Democratic Republic of the Congo", on the DRC national cumulative case
and death series.

## Contents

- `extracted_epi_figures.json`: the extracted, source-attributed case and death figures (the input data).
- `official_cases.csv`, `official_deaths.csv`: the official national series (the validation target).
- `reconcile.py`: reconstructs the cumulative curve from the figures using a tier weighting (official > wire > local > aggregator), then weighted isotonic regression.
- `nowcast.py`: vintage (no-look-ahead) reconstruction, lag estimation and the backtested nowcast.
- `validate.py`: the reconstructed curve against the official series (validation table).
- `benchmark.py`: the no-look-ahead backtest, open-source nowcast vs projecting the last official figure forward, median error by horizon with a one-sided paired Wilcoxon test.
- `phase_transition.py`: median error by epidemic movement (steady, moderate, turning point), reproducing the result that the gain concentrates at the turning points, where extrapolating the official trend is furthest off.
- `deployment.py`: reporting-lag characterisation and real-time deployment, using the live snapshots below and the publication dates.

### Data snapshots
- `official_cases.csv`, `official_deaths.csv`, `extracted_epi_figures.json`: the frozen validation window (to 3 August), for `benchmark.py`, `phase_transition.py` and `validate.py`.
- `official_cases_live.csv`, `official_deaths_live.csv` (to 24 August), `extracted_epi_figures_live.json` (open-source figures to 27 August) and `insp_publication_dates.json` (each observation date mapped to the day its situation report was published, from the source repository's commit history), for `deployment.py`.

## Run

```
python3 benchmark.py         # nowcast vs official-only by horizon
python3 phase_transition.py  # error by epidemic movement (the turning-point result)
python3 deployment.py        # reporting lag and real-time deployment to late August
python3 validate.py          # reconstructed curve vs official (validation table)
python3 nowcast.py           # backtested nowcast (draws a plot if matplotlib is present)
```

Requires `numpy` and `scipy`. `nowcast.py` additionally uses `matplotlib` for its plot and skips the plot if it is not installed. The publication dates can be regenerated from the public data repository's commit history (each situation report is committed on the day it is published); the mapping is included here for convenience.

## What is not included, and why

Two production components are not included:

1. The step that turns source text into the structured figures in `extracted_epi_figures.json`. Only its output, the figures, is shared.
2. The more detailed source weighting used for the paper's headline numbers. This bundle uses the simpler, fully specified tier weighting instead.

This tier version reaches the same conclusion, that the open-source nowcast beats projecting the last official figure forward once the official figure is a few days stale, with slightly larger errors, chiefly on deaths. The direction and significance of the result are reproducible here.

## Scope

DRC national cumulative confirmed cases and deaths, established phase (official count at or above 100). One outbreak. The ground truth is the eventual official figure, so the method is validated as being faster to that figure, not more accurate than it.
