## Supplementary material for "Nowcasting outbreak case and death counts from open-source reporting A validation on the 2026 Ebola (Bundibugyo) outbreak in the Democratic Republic of the Congo": Reproducibility bundle: extracted figures, official series, and backtest code: reconcile.py

```
"""Tier-weighted cumulative epi-curve reconstruction: the simple, fully specified
version referenced in the paper's data and code availability statement. It reproduces
the paper's central conclusion (the open-source nowcast beats projecting the last
official figure forward in the reporting gap). Two production components are not
included: the step that extracts the structured figures from source text (only its
output, extracted_epi_figures.json, is shared), and the more detailed source weighting
used for the paper's headline numbers. This tier version reaches the same conclusion.
"""
import os, json
from collections import defaultdict

HERE = os.path.dirname(__file__)
IN = os.path.join(HERE, "extracted_epi_figures.json")
OUTBREAK_START = "2026-05-01"   # exclude the earlier outbreak

TIER_W = {"official": 4.0, "wire": 3.0, "local": 2.0, "aggregator": 1.0, "other": 1.0}
TIER_RANK = {"official": 4, "wire": 3, "local": 2, "aggregator": 1, "other": 0}
NATIONAL = {"national_drc"}     # DRC-only estimand, to match the official series

OFFICIAL = ["who", "world health", "oms", "africa cdc", "cdc", "ministry", "ministère", "ministere",
            "santé", "sante", "unicef", "ocha", "reliefweb", "united nations", "un news"]
WIRE = ["reuters", "associated press", " ap ", "afp", "agence france", "bbc", "france 24", "france24",
        "deutsche welle", "dw ", "al jazeera", "aljazeera", "anadolu", "guardian", "cnn", "xinhua",
        "sana", "yahoo", "abc news", "cbs", "nbc", "npr", "financial times", "economist"]
LOCAL = ["radio okapi", "actualite", "actualité", "mediacongo", "7sur7", "prosperite", "prospérité",
         "kivu", "daily monitor", "new vision", "monitor", "allafrica", "congo", "kampala", "rdc"]
AGGREGATOR = ["google news"]


def tier(source):
    s = (source or "").lower()
    if any(k in s for k in AGGREGATOR):
        return "aggregator"
    if any(k in s for k in OFFICIAL):
        return "official"
    if any(k in s for k in WIRE):
        return "wire"
    if any(k in s for k in LOCAL):
        return "local"
    return "other"


def isotonic(xs, ys, ws):
    """Weighted isotonic regression (non-decreasing), pool-adjacent-violators."""
    n = len(ys)
    if n == 0:
        return []
    v_stack, w_stack, cnt_stack = [], [], []
    for i in range(n):
        v, w, c = float(ys[i]), float(ws[i]), 1
        while v_stack and v_stack[-1] > v:
            pv, pw, pc = v_stack.pop(), w_stack.pop(), cnt_stack.pop()
            v = (pv * pw + v * w) / (pw + w); w = pw + w; c = pc + c
        v_stack.append(v); w_stack.append(w); cnt_stack.append(c)
    out = []
    for v, c in zip(v_stack, cnt_stack):
        out.extend([v] * c)
    return out


def obs_date(f):
    d = f.get("as_of")
    if isinstance(d, str) and len(d) == 10 and d[:4] == "2026":
        return d
    return f.get("date")


def load():
    figs = json.load(open(IN, encoding="utf-8"))
    keep = []
    for f in figs:
        if f.get("outbreak") is not True:
            continue
        if not isinstance(f.get("value"), int) or f["value"] <= 0:
            continue
        d = obs_date(f)
        if not d or d < OUTBREAK_START:
            continue
        f["_date"] = d
        f["_pubdate"] = f.get("date")
        f["_tier"] = tier(f.get("source"))
        if f["_tier"] == "official":
            continue                          # exclude agency republications: use no official data
        keep.append(f)
    return keep


def reconcile_metric(figs, metric):
    """Sorted [(date, reconciled_cumulative, min, max, n)]. Tier-preferred median per
    day (official tier if present, else best tier available; median within that tier),
    then weighted isotonic to enforce a non-decreasing cumulative curve."""
    wanted = {metric} if isinstance(metric, str) else set(metric)
    rows = [f for f in figs if f.get("metric") in wanted
            and f.get("count_type") == "cumulative" and f.get("geography") in NATIONAL]
    by_date = defaultdict(list)
    for f in rows:
        by_date[f["_date"]].append(f)
    dates = sorted(by_date)
    xs, ys, ws, lo, hi, ns = [], [], [], [], [], []
    for d in dates:
        group = by_date[d]; allv = [g["value"] for g in group]
        best = max(TIER_RANK[g["_tier"]] for g in group)
        top = sorted(g["value"] for g in group if TIER_RANK[g["_tier"]] == best)
        rep = top[len(top) // 2]
        w = TIER_W[[t for t, r in TIER_RANK.items() if r == best][0]]
        xs.append(d); ys.append(rep); ws.append(w)
        lo.append(min(allv)); hi.append(max(allv)); ns.append(len(group))
    fitted = isotonic(xs, ys, ws)
    return list(zip(xs, [round(v) for v in fitted], lo, hi, ns))
```
