## Supplementary material for "Nowcasting outbreak case and death counts from open-source reporting A validation on the 2026 Ebola (Bundibugyo) outbreak in the Democratic Republic of the Congo": Reproducibility bundle: extracted figures, official series, and backtest code: validate.py

```
"""Validate the reconstructed open-source curve against the official series (national
cumulative confirmed cases and deaths). The reconstruction never saw the official
line-list: how close does fused open-source reporting land, and does it lead or lag?

Run:  python3 validate.py
"""
import os, csv

HERE = os.path.dirname(__file__)


def load_official(fname, col):
    out = {}
    with open(os.path.join(HERE, fname), encoding="utf-8") as fh:
        for row in csv.DictReader(fh):
            if row.get("nom") == "DRC" and row.get("date") and row.get(col):
                out[row["date"]] = int(float(row[col]))
    return out


def interp(series, d):
    """Official cumulative value on date d (carry-forward last known)."""
    ks = [k for k in sorted(series) if k <= d]
    return series[ks[-1]] if ks else None


def _stats(pairs):
    errs = sorted((v - o) / o for _, v, o in pairs)
    ape = sorted(abs(e) for e in errs)
    n = len(pairs)
    med = ape[n // 2]
    medbias = errs[n // 2]
    return n, 100 * med, 100 * medbias, 100 * sum(a <= 0.10 for a in ape) / n


def compare(recon, official, label, established=100):
    rows = [(p["date"], p["cumulative"]) for p in recon]
    pairs = [(d, v, interp(official, d)) for d, v in rows]
    pairs = [(d, v, o) for d, v, o in pairs if o]
    if not pairs:
        print(f"{label}: no overlap"); return
    early = [p for p in pairs if p[2] < established]
    est = [p for p in pairs if p[2] >= established]
    print(f"\n=== {label} vs official ({len(pairs)} overlapping dates) ===")
    print("  (median metrics, robust to tiny early-outbreak denominators)")
    n, med, mb, w10 = _stats(pairs)
    print(f"  ALL          n={n:3d}  median APE {med:5.1f}%   median bias {mb:+5.1f}%   within +/-10%: {w10:3.0f}%")
    if early:
        n, med, mb, w10 = _stats(early)
        print(f"  EARLY <{established:<4d} n={n:3d}  median APE {med:5.1f}%   median bias {mb:+5.1f}%   (OSINT catches suspected/rumour before official confirms)")
    if est:
        n, med, mb, w10 = _stats(est)
        print(f"  ESTABLISHED  n={n:3d}  median APE {med:5.1f}%   median bias {mb:+5.1f}%   within +/-10%: {w10:3.0f}%")
    for d, v, o in pairs[-1:] + [p for p in pairs if p[0] in ("2026-06-01", "2026-06-15", "2026-07-01", "2026-07-15")]:
        print(f"    {d}  reconstructed={v:5d}  official={o:5d}  ({100*(v-o)/o:+.0f}%)")


def main():
    from reconcile import load, reconcile_metric
    figs = load()
    off_cases = load_official("official_cases.csv", "national_cumulative_confirmed_cases")
    off_deaths = load_official("official_deaths.csv", "national_cumulative_confirmed_deaths")
    print(f"official cases {min(off_cases)}..{max(off_cases)} "
          f"({off_cases[max(off_cases)]}), deaths ({off_deaths[max(off_deaths)]})")
    cases = [{"date": d, "cumulative": v} for d, v, *_ in reconcile_metric(figs, {"confirmed_cases", "total_cases"})]
    deaths = [{"date": d, "cumulative": v} for d, v, *_ in reconcile_metric(figs, {"deaths"})]
    compare(cases, off_cases, "cases")
    compare(deaths, off_deaths, "deaths")


if __name__ == "__main__":
    main()
```
